# Cortisol Stress Response is Associated with Iron Status in Pregnancy

**DOI:** 10.64898/2026.06.22.26356237

**Authors:** Brie M. Reid, Georgia F. Celestin, Michael K. Georgieff, Kimberley Mbayiwa, Kate Keenan

## Abstract

**Background:** Iron deficiency (ID) affects up to 40% of pregnant women in the third trimester, even in highly resourced and iron-supplemented populations, with adverse consequences for maternal health and long-term offspring development. Psychological stress may compromise iron status through hypothalamic-pituitary-adrenocortical (HPA) axis dysregulation and inflammation, but no study has directly examined cortisol in relation to iron status across human pregnancy.

**Objective:** This longitudinal study examined associations between HPA function and maternal iron status across pregnancy and tested whether IL-6 and CRP mediated the relationship between cortisol and ferritin across gestation.

**Methods:** One hundred sixty-eight pregnant Black women with Medicaid insurance completed up to four laboratory assessments across pregnancy. Salivary cortisol was measured before and in response to the Trier Social Stress Test, yielding basal and reactive cortisol indices. Serum ferritin, IL-6, and CRP were collected at each visit. Trimester-specific regression models examined cortisol reactivity in relation to ferritin; linear mixed-effects models with moderated mediation tested whether basal cortisol predicted ferritin via inflammation.

**Results:** Higher cortisol reactivity was associated with lower ferritin specifically in the third trimester (std. β = −0.197, p = .004). Higher basal cortisol predicted a steeper IL-6 rise across gestation (*p* = .002), and IL-6 was positively associated with ferritin (b = 0.236, p = .006), consistent with inflammatory iron sequestration. The indirect effect of basal cortisol on ferritin via IL-6 was statistically significant, and higher basal cortisol was negatively associated with cortisol reactivity in the third trimester. No pathway was observed through CRP.

**Conclusion:** Greater cortisol reactivity predicted lower third-trimester ferritin, a pattern that suggests cumulative iron depletion, atypically sustained HPA reactivity in late pregnancy, or both. To our knowledge, this is the first prospective study linking cortisol reactivity to iron status across human pregnancy, identifying maternal stress physiology as a novel target for understanding and addressing gestational iron deficiency.

## INTRODUCTION

Iron deficiency (ID) is highly prevalent during pregnancy, affecting an estimated 30–40% of pregnant women globally (1,2), and carrying significant consequences for maternal health and offspring development (3–6), including increased maternal morbidity and mortality (reviewed in (7)). ID is not confined to populations with inadequate intake: a recent longitudinal study found that over 51% of primiparous, non-anemic women in Ireland were iron deficient (ferritin < 15 μg/L) by the third trimester, despite most taking oral iron supplements. Iron requirements increase by nearly seven-fold across pregnancy due to maternal blood volume expansion, placental iron demand, and fetal iron accrual. Accordingly, the frequent failure to meet this demand makes maternal iron deficiency the leading cause of reduced fetal iron supply (4,8). Ferritin concentrations >60 μg/L in early pregnancy have been identified as protective against the risk of later gestational iron deficiency (ferritin < 15 μg/L in the third trimester), yet these levels are challenging to maintain (9). Maternal ferritin at delivery below 13.4 μg/L compromises neonatal iron stores (10), and insufficient maternal-fetal iron transfer is associated with a range of neurocognitive and mental health disorders in offspring with consequences for later education attainment, occupational outcomes, and wellbeing (3–6).

Populations that face disproportionate stress burden are also those at higher risk of iron deficiency: ID is more prevalent among pregnant women in low- and middle-income countries and among those with fewer socioeconomic resources in high income countries (2,11,12). In the United States, one in five women experience more than one stressful life event during pregnancy, with higher rates among non-Hispanic Black women and American Indian/Alaska Native women (6,13). The overlap between populations at risk for iron deficiency and those experiencing elevated stress suggests a potential causal link, supported by a growing body of animal and human evidence. In rodent models, psychological stress reduces serum iron and hemoglobin relative to non-stressed controls (14), and experimental prenatal stress in non-human primates exacerbates iron deficiency in offspring during periods of high iron demand (15). In humans, acute stress disrupts iron status in adults following sustained stress exposure (16), and severe family conflict stress in the first year of life is associated with lower infant iron status even in healthy, full-term infants (17). During pregnancy, both objective and subjective stress measures are associated with lower cord blood ferritin and offspring iron status (18–20), and recent work found that self-reported stress predicted worse fetal iron transfer (21). Despite convergent evidence linking prenatal stress to offspring iron outcomes, the mechanisms by which maternal stress affects maternal iron status during pregnancy remain poorly understood (22).

One plausible biological pathway runs through the hypothalamic-pituitary-adrenal (HPA) axis. Psychosocial stress activates the HPA axis, triggering cortisol release and engaging immune-inflammatory pathways through crosstalk between the stress and immune systems (23). When chronic, stress can dysregulate HPA function and impair cortisol’s regulatory effects on immune activity, driving a sustained pro-inflammatory response (24). In this way, stress may impair iron uptake even when dietary intake is sufficient. Interleukin-6 (IL-6), a pro-inflammatory cytokine that is elevated in response to both psychosocial and infectious stress (22), may play a central role in this pathway. IL-6 stimulates the hepatic production of C-reactive protein (CRP) and upregulates hepcidin, a hormone that suppresses dietary iron absorption in the gut and may also impair maternal-fetal iron transfer (8,25). In pregnancy, hepcidin concentrations are normally suppressed to facilitate sufficient iron transfer to the fetus via the placenta (4,6,8). Stress-driven upregulation of IL-6 may therefore interfere with this adaptive suppression, reducing iron bioavailability at precisely the time when maternal and fetal iron demand is greatest. Consistent with this hypothesis, high stress and low social support during pregnancy are associated with elevated IL-6 and L-1β across trimesters (26), and pro-inflammatory responses to cortisol have been linked to impaired iron absorption in non-pregnant populations (6,27,28). Together, these mechanisms support the hypothesis that stress in pregnancy compromises iron status even in the absence of infection or dietary inadequacy, possibly through an HPA-immune axis that elevates IL-6 and ultimately reduces iron bioavailability (6).

Two biologically distinct aspects of HPA function may be relevant to iron metabolism across pregnancy. First, basal cortisol rises across gestation to reach circulating concentrations roughly seven times higher than the non-pregnant state, driven in part by placental corticotropin-releasing hormone and reduced sensitivity to glucocorticoid feedback (29). This normative rise may interact with stress-driven inflammatory signaling to progressively alter iron metabolism across gestation. Second, cortisol reactivity to acute stress tends to attenuate across pregnancy, particularly in the third trimester, interpreted as a physiological adaptation that shields both the mother and the fetus from prolonged glucocorticoid exposure (30–32). Failing to show this expected attenuation of cortisol reactivity in late pregnancy may be one way stress physiology can influence iron status. These two aspects of HPA function may therefore have distinct consequences for iron status across pregnancy, motivating the examination of both basal and reactivity cortisol indices in the present study.

To our knowledge, no prior study has directly examined whether cortisol, measured at resting or reactively, is associated with maternal iron status across human pregnancy. The present study addresses this gap in our understanding of how stress may impact iron status using a prospective longitudinal design in a sample of pregnant Black women living in stressed urban environments. We examined two distinct indices of HPA function (cortisol reactivity to acute stress and basal cortisol across gestation) in relation to serum ferritin and tested whether IL-6 and CRP mediated these associations. We hypothesized that (1) greater cortisol reactivity would be associated with lower ferritin, particularly in the third trimester when fetal iron demand is greatest and maternal reserves are most depleted; (2) higher basal cortisol rising across gestation would be associated with lower ferritin over time; and (3) inflammatory markers — specifically IL-6 and CRP — would mediate the association between cortisol and ferritin, consistent with an HPA-immune pathway linking stress physiology to iron status across pregnancy.

## METHOD

### Participants

Participants were recruited as part of the Nutrition and Pregnancy Study (NAPS: NCT02647723), a double-blind randomized controlled trial of fatty acid supplementation for Black women living in Chicago, IL (*n* = 168; see Table 1 for demographic information; Keenan et al., 2016 for detailed methods) (33). Participants met inclusion criteria for the study if they were pregnant and between 10-18 weeks of gestation, self-identified as Black, between 18 and 34 years, received public assistance i.e., Medicaid insurance, and reported consuming less than two servings of sea fish weekly. Exclusion criteria included serious medical complications, regular use of steroid medications, blood thinners, or anti-coagulants, use of psychotropic medications, substance use, and allergy to fish, soy, strawberries, or iodine. A participant flow diagram is shown in Supplemental Figure 1. All procedures for this study were approved by the University of Chicago Institutional Review Board. Participants provided written informed consent before initiation of study procedures.

**TABLE 1.** Demographics.

|  |  |  |  |  |
| --- | --- | --- | --- | --- |
| Maternal Age (Visit 1) |  |  |  |  |
| Mean (SD) | 25.69 (4.21) |  |  |  |
| Maternal BMI (earliest) |  |  |  |  |
| Mean (SD) | 32.04 (9.51) |  |  |  |
| Gestational Age at BMI |  |  |  |  |
| Mean (SD) | 26.84 (12.9) |  |  |  |
|  | Trimester 1 | Trimester 2 | Trimester 3 | p-value |
| Gestational Age (weeks) |  |  |  |  |
| Mean (SD) | 12.00 (± 1.24) | 18.97 (± 1.92) | 31.76 (± 1.84) | - |
| Cortisol Response (ln) |  |  |  |  |
| Mean (SD) | -0.01 (± 0.18) | -0.05 (± 0.19) | -0.04 (± 0.09) | 0.228 |
| Missing | 1 (1.3%) | 10 (6.5%) | 33 (22.8%) |  |
| BRINDA Serum Ferritin (ug/L) |  |  |  |  |
| Mean (SD) | 94.97 (± 57.21) | 75.19 (± 48.28) | 48.85 (± 41.50) | < 0.001*** |
| Missing | 10 (12.5%) | 17 (11.0%) | 33 (22.8%) |  |
| Raw Serum Ferritin (ug/L) |  |  |  |  |
| Mean (SD) | 89.18 (± 53.48) | 76.60 (± 48.81) | 55.05 (± 45.33) | < 0.001*** |
| Missing | 10 (12.5%) | 17 (11.0%) | 33 (22.8%) |  |
| Perceived Stress Scale |  |  |  |  |
| Mean (SD) | 14.94 (± 8.07) | 14.76 (± 7.52) | 13.48 (± 7.41) | < 0.001*** |
| Missing | 1 (1.3%) | 3 (1.9%) | 9 (6.2%) |  |
| CRP (mg/L) |  |  |  |  |
| Mean (SD) | 9.00 (± 9.99) | 11.78 (± 12.15) | 10.12 (± 10.70) | 0.002** |
| Missing | 2 (2.5%) | 6 (3.9%) | 20 (13.8%) |  |
| IL-6 (fg/mL) |  |  |  |  |
| Mean (SD) | 2207.88 (± 1854.34) | 2647.97 (± 2405.23) | 2698.48 (± 1928.35) | 0.0521+ |
| Missing | 2 (2.5%) | 6 (3.9%) | 20 (13.8%) |  |
| HB (g/dL) |  |  |  |  |
| Mean (SD) | 11.85 (± 1.17) | 10.98 (± 0.95)<sup>†</sup> | 10.80 (± 1.34) | < 0.001*** |
| Missing | 47 (58.8%) | 117 (75.5%) | 40 (27.6%) |  |

Note. <sup>+</sup> $p < .10$ , \*\* $p < .01$ , \*\*\* $p < .001$ ; For participants that had multiple visits to the lab within a single trimester, the participant's data were averaged within that trimester. <sup>†</sup> Note: in the second trimester only, participants in the placebo group had higher levels of serum hemoglobin than participants in the DHA group ( $t = 2.123$ , $p = .041$ ).

### Procedure

Prenatal visits occurred between April 2016 and June 2021. Participants completed four lab visits across pregnancy between 12:00–5:00 pm to account for diurnal cortisol variation, with a one-hour fast required prior to each visit. Each visit included an acclimation period, a standardized laboratory stressor to assess acute cortisol reactivity, a recovery period during which participants completed demographic and psychosocial questionnaires, and a venous blood draw from which ferritin and inflammatory markers were derived. Saliva was collected throughout as described below. Blood was processed immediately post-visit; serum and saliva were stored at −80°C. BMI was extracted from medical charts (n = 121); the earliest available measurement was used as a proxy for pre-pregnancy BMI; for most participants this was measured in the third trimester (n = 71). All 168 participants completed visit 1; 144, 134, and 129 completed visits 2, 3, and 4, respectively, with attrition primarily due to the COVID-19 pandemic and missed or canceled visits.

### Salivary Cortisol

The Trier Social Stress Test (TSST) is a validated laboratory protocol for inducing moderate psychological stress and measuring cortisol reactivity (34) that has been used successfully in pregnant populations (30,35–41). The specific protocol used in the present study has been described previously (37). Start time was recorded to account for diurnal variation. Five saliva samples were collected via 2-minute mouth swabs by trained researchers at: lab arrival, at 20 minutes after arrival, pre-TSST, and 20, 40, and 50 minutes post-TSST. Frozen samples were transported to the Clinical Research Center at the University of Chicago, where salivary cortisol was quantified using the Salimetrics HS Salivary Cortisol EIA kit. Inter-and intra-assay coefficients of variation were 3.9% and 6.7% at high concentrations and 7.1% and 6.9% at low concentrations, respectively. Cortisol data were missing for 5, 11, 20, and 30 participants at visits 1–4, respectively.

### Self-reported stress

Self-reported stress was measured using the Perceived Stress Scale (PSS) (42). The PSS is a 10-item self-report measure that assesses participants’ feelings regarding stressors and their controllability over the past month using a 5-point Likert scale (sum score range of 0-40). The PSS has good internal consistency and test-retest reliability (43). The PSS was missing for 2 participants at visit 1, 0 participants at visit 2, 0 participants at visit 3, and 2 participants at visit 4.

### Iron Status

Frozen serum samples were transported to the Clinical Research Center at the University of Chicago, where serum ferritin was quantified using the Sigma-Aldrich Human Ferritin ELISA Kit. Ferritin data were missing for 19, 24, 30, and 33 participants at visits 1–4, respectively. Hemoglobin values were extracted from participants’ medical charts and matched to the nearest research visit using gestational age at medical appointment. Hemoglobin data were available for 130 participants: 61 had data in one trimester, 49 in two trimesters, and 20 in all three, yielding 59, 43, and 117 observations in trimesters 1, 2, and 3, respectively.

### Inflammation

Frozen serum samples were shipped to the Central Ligand Assay Satellite Services (CLASS) Laboratory at the University of Michigan School of Public Health for analysis. CRP and IL-6 were quantified via enzyme-linked immunosorbent assay (ELISA), with coefficients of variation <5% for both analytes. Missing data were minimal: CRP was missing for 6, 5, 9, and 15 participants at visits 1–4 respectively, and IL-6 for 6, 6, 8, and 16 participants.

### Statistical Analysis

Serum ferritin concentrations were adjusted for inflammation using the Biomarkers Reflecting Inflammation and Nutritional Determinants of Anemia (BRINDA) method and corresponding R package (44), which corrects ferritin concentrations using CRP and/or alpha-1-acid glycoprotein (AGP) to improve interpretability in the presence of inflammation (45,46). Consistent with BRINDA recommendations, internal lowest-decile CRP concentrations were used for adjustment at each visit, as our sample’s internal deciles differed substantially from the BRINDA reference deciles developed for non-pregnant women of childbearing age. This approach has been validated in pregnant and non-pregnant populations and consistently increases estimates of iron deficiency prevalence after accounting for inflammation (46–48).

All analyses were conducted in R (Version 2024.12.1+563). Iron deficiency was defined using standard pregnancy thresholds: ferritin <15 μg/L (iron deficient), 15–30 μg/L (at risk), and >30 μg/L (iron sufficient) (8,49). Ferritin and cortisol reactivity values were log-transformed. The sample collected 20 minutes after arrival was used to measure basal cortisol. Cortisol reactivity was calculated as the difference between basal levels and levels measured at 20 minutes post TSST (when peak response to a stressor is likely to be measurable in saliva). CRP, IL-6, and hemoglobin were visually inspected for outliers and implausible values; CRP and IL-6 were subsequently log-transformed. No participants were excluded for incomplete or implausible iron or inflammation data.

To assess the impact of cortisol reactivity on ferritin in each trimester, we regressed log-transformed ferritin on log-transformed cortisol reactivity, controlling for gestational age and RCT group assignment. Visit data were categorized by trimester using continuous gestational age: first trimester (<14 weeks), second trimester (14–26 weeks), and third trimester (>26 weeks). For trimester 1 (N = 80), a single linear model was used. For trimesters 2 and 3, where most participants contributed multiple visits (N = 107 and N = 103, respectively), linear mixed-effects models were fit using the lme4 package (50), with a random effect at the subject level to account for repeated measures.

To examine whether basal cortisol was associated with inflammatory markers (CRP, IL-6) across gestation, and whether those markers in turn predicted ferritin, basal (pre-TSST), we fit three sets of multilevel models: (1) a basal cortisol (pre-TSST) × gestational age interaction on ferritin to characterize the overall cortisol–ferritin association across pregnancy; (2) Path A models regressing the cortisol × gestational age interaction onto CRP and IL-6 separately; and (3) Path B models regressing CRP and IL-6 separately onto ferritin, retaining the cortisol × gestational age interaction term. Indirect effects were estimated at five gestational age windows (12, 20, 28, 34, and 38 weeks) using Monte Carlo simulation (k = 20,000). Gestational age was centered at 28 weeks to aid interpretability around third trimester onset. As inflammatory variables were included as predictors, ferritin values in these models were not inflammation-corrected to avoid overcorrection.

Sensitivity analyses examined maternal age and time of day of cortisol collection as additional covariates in the main models and assessed whether concurrent self-reported stress (PSS total score) accounted for the cortisol reactivity–ferritin association. Additional sensitivity analyses were conducted to test whether cortisol reactivity and ferritin differed by RCT group or BMI category (≥30 vs. <30) using independent samples t-tests, whether cortisol reactivity and hemoglobin were significantly correlated, and whether cortisol reactivity predicted CRP or IL-6 after controlling for gestational age.

## RESULTS

Demographic information is presented in Table 1. Serum ferritin declined significantly across pregnancy, F(2, 364.7) = 61.066, *p* < .001, with significant differences in iron status risk classifications across trimesters, χ²(6) = 41.977, *p* < .001. Using a threshold of <15 μg/L, iron deficiency rates ranged from 7.1% to 24.1%, peaking in the third trimester (Figure 1). Anemia rates (hemoglobin <11 g/dL) ranged from 21.2% to 55.2%, likewise peaking in the third trimester. Iron deficiency anemia (ferritin <30 μg/L and hemoglobin <11 g/dL) was rare in the first and second trimesters (1.5% and 0.9%, respectively) but reached 17.9% by the third trimester. Gestational age at birth was not associated with maternal ferritin in the first trimester (*r* = -0.028, *p* = 0.0828), second trimester (*r* = 0.017, *p* = 0.847), or third trimester (*r* = -0.171, *p* = 0.073). Self-reported stress did not significantly predict ferritin in any trimester (trimester 1: *p* = .164; trimester 2: *p* = .061; trimester 3: *p* = .926). Intercorrelations among study variables are provided in Supplementary Tables 1–3.

**Figure 1.**
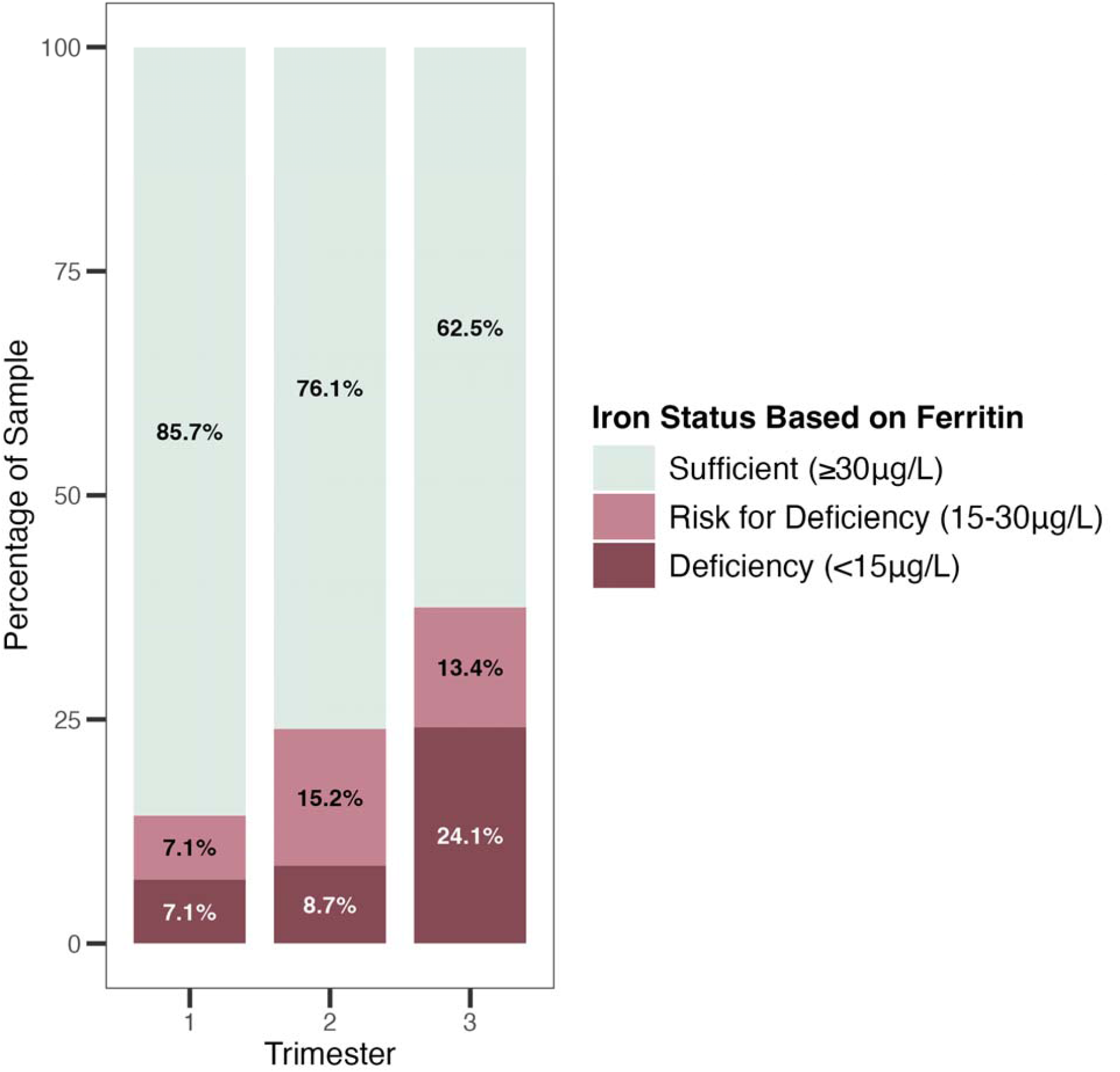
Iron status classification based on ferritin levels across trimesters shows increasing rates of iron deficiency as pregnancy progresses.

### Cortisol and Ferritin

Cortisol reactivity to the TSST was negatively associated with serum ferritin in the third trimester (β = −0.197, 95% CI [−0.329, −0.064], *p* = .004; Figure 3); no significant associations were observed in the first or second trimesters (Table 2). This association remained significant after controlling for maternal age and time of day of cortisol collection (β = −0.186, 95% CI [−0.322, −0.050], *p* = .008), and concurrent self-reported stress (β = −0.197, 95% CI [−0.330, −0.063], *p* = .004), none of which were significant predictors in any model. No significant differences in cortisol reactivity or serum ferritin were found by RCT group in any trimester. See supplemental information for hemoglobin by RCT group across trimesters.

**Figure 2.**
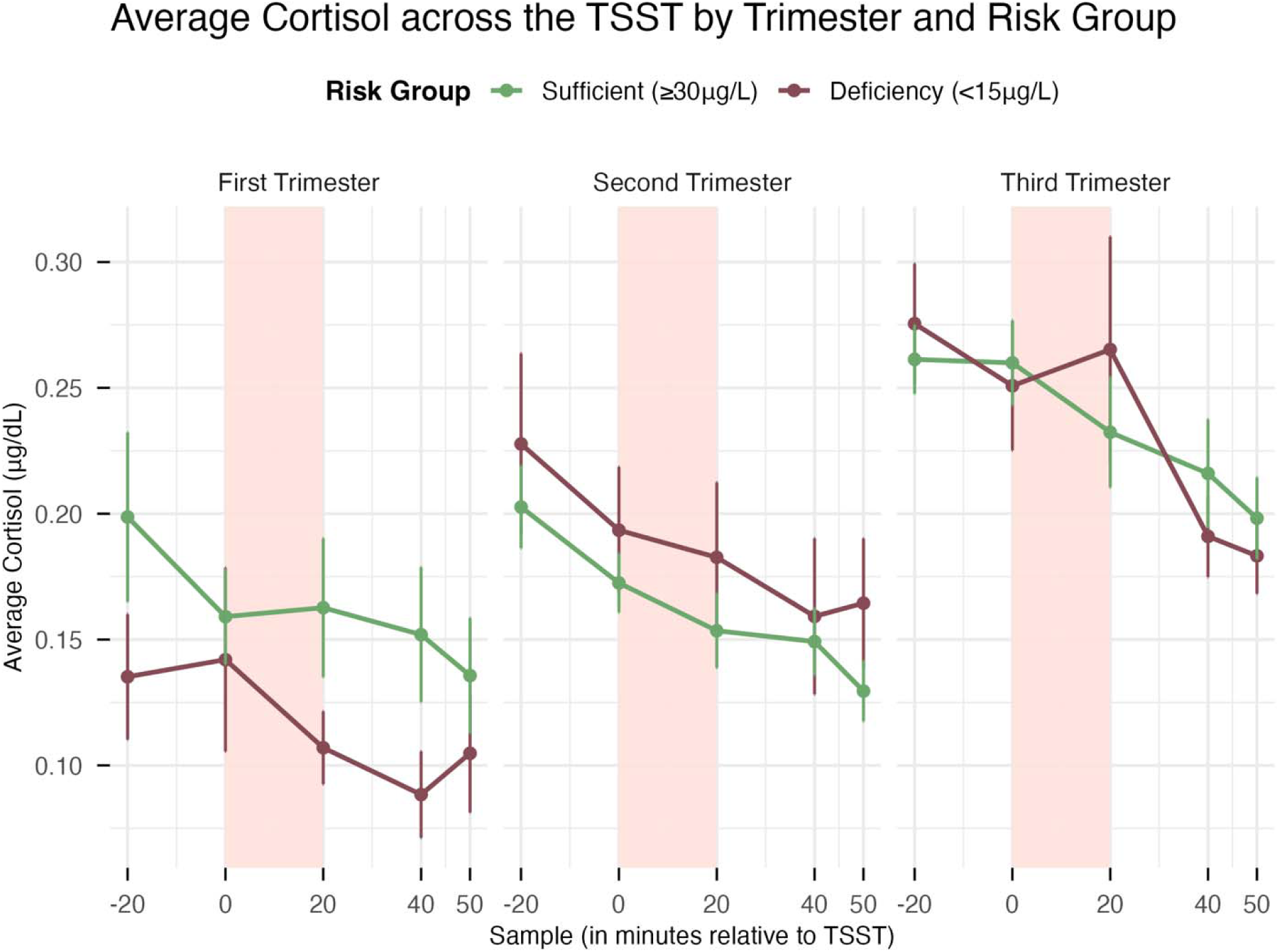
Higher Cortisol Reactivity Predicts Lower Iron Stores in the Third Trimester. Cortisol reactivity to the Trier Social Stress Test (TSST) across pregnancy trimesters, stratified by iron status (iron deficient vs. iron sufficient). This figure is for descriptive purposes only and does not reflect the analytical model. In statistical models, a significant negative association between cortisol reactivity and ferritin was observed in the third trimester only, such that greater cortisol reactivity was associated with lower ferritin levels. Across trimesters, mean cortisol levels declined from –20 to 50 minutes post-TSST: first trimester (M = 0.178 → 0.120, t(2383) = 2.54, p = .011); second trimester (M = 0.213 → 0.132, t(2387) = 6.17, p < .001); third trimester (M = 0.276 → 0.201, t(2384) = 5.09, p < .001).

**Figure 3.**
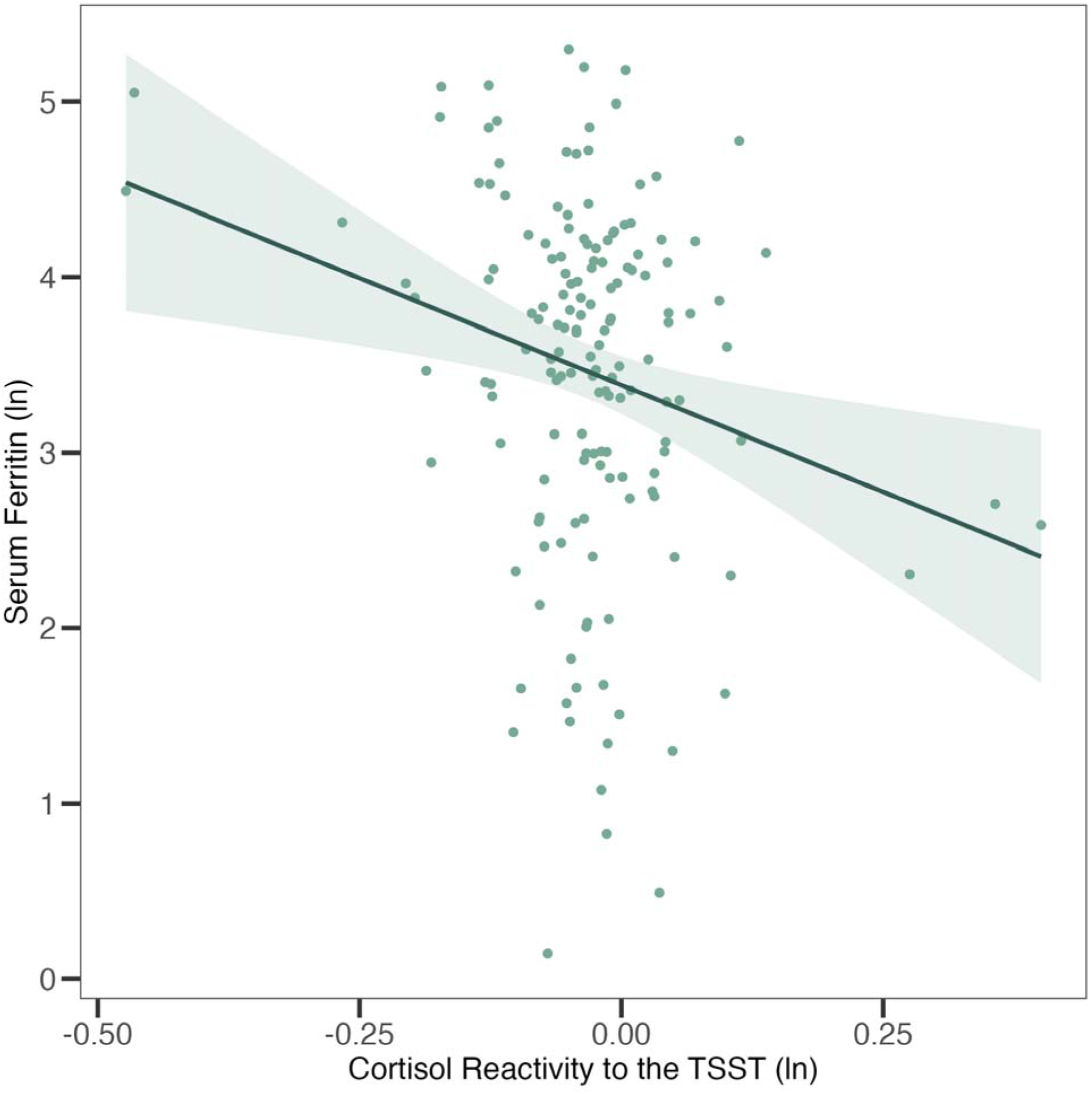
Serum ferritin levels are negatively associated with elevated cortisol reactivity. Association between cortisol reactivity to the Trier Social Stress Test (TSST) and serum ferritin in the third trimester. Both variables are natural log-transformed. The regression line and 95% confidence interval are shown. Higher cortisol reactivity was associated with lower serum ferritin (β = −0.197, 95% CI [−0.329, −0.064], *p* = .004), controlling for gestational age and RCT group assignment.

**TABLE 2.** Linear Regression Output of Salivary Cortisol and Serum Ferritin Across Pregnancy.

| *Serum Ferritin Across Pregnancy* | *b* | *SE* | Std. $\beta$ | *t* | *95% CI* | *p* |
| --- | --- | --- | --- | --- | --- | --- |
| Salivary Cort. Reactivity Trimester 1 | 0.228 | 0.559 | 0.050 | 0.408 | -1.067, 1.167 | 0.684 |
| Salivary Cort. Reactivity Trimester 2 | -0.110 | 0.426 | -0.015 | -0.257 | -0.130, 0.100 | 0.797 |
| Salivary Cort. Reactivity Trimester 3 | -2.072 | 0.708 | -0.197 | -2.926 | -0.329, -0.064 | **0.004\*\*** |

*Note.* \*\* $p < .01$

Cortisol reactivity was not associated with hemoglobin in any trimester 1 (*r* = −0.076, *p* = .676, *r* = −0.108, *p* = .536, and *r* = 0.104, *p* = .350, for trimesters 1, 2, and 3, respectively). Third trimester cortisol reactivity and ferritin did not differ significantly between participants with BMI ≥ 30 and BMI < 30 at earliest BMI measurement available (cortisol reactivity: *M* = −0.041 vs. −0.034, *t*(67.8) = 0.307, *p* = .760; ferritin: *M* = 3.521 vs. 3.287, *t*(76.9) = −0.955, *p* = .343). Figure 2 depicts mean salivary cortisol levels (μg/dL) across the TSST by trimester, stratified by iron-sufficient (≥30 μg/L) and iron-deficient (<15 μg/L) status.

As ferritin and cortisol reactivity were measured concurrently, we tested for temporal precedence among participants with two third-trimester visits (*n* = 103), controlling for gestational age at outcome assessment. Earlier cortisol reactivity did not significantly predict later ferritin (β = −0.150, 95% CI [-0.399, 0.099], *p* = .232), though the effect direction was consistent with our primary findings. Earlier ferritin likewise did not predict later cortisol reactivity (β = 0.071, 95% CI [-0.183, 0.325], *p* = .578).

There was a marginally significant interaction between basal cortisol and gestational age predicting ferritin (*b* = 0.015, *SE* = 0.009, *t*(328) = 1.693, *p* = .091, std. β = 0.066), suggesting a trend toward a stronger cortisol–ferritin association with advancing gestation, though this did not reach significance. We then examined whether basal cortisol (within-person, trimester-level mean) predicted BRINDA-corrected ferritin (within-person, trimester-level mean) using three separate linear models (one per trimester). Basal cortisol was not a significant predictor of ferritin in trimester 1 (β = 0.024, 95% CI [−0.220, 0.268], *p* = .845), trimester 2 (β = −0.031, 95% CI [−0.205, 0.143], *p* = .725), or trimester 3 (β = 0.029, 95% CI [−0.170, 0.228], *p* = .774).

We conducted a post hoc analysis to contextualize third trimester basal and reactivity findings and examined whether basal cortisol (within-person, trimester-level mean) was associated with third trimester cortisol reactivity (within-person mean) across pregnancy. Using three separate linear models (one per trimester), we found a negative association only between third trimester cortisol reactivity and third trimester basal cortisol (β = −0.323, 95% CI [−0.502, −0.145], *p* = .001), suggesting that mothers with higher basal cortisol tended to show lower cortisol reactivity by the third trimester. There were no significant associations with third trimester cortisol reactivity and basal cortisol in the first (β = −0.011, 95% CI [−0.311, 0.290], *p* = .943) or second trimesters (β = −0.111, 95% CI [−0.301, 0.079], *p* = .251).

### Cortisol, Inflammation, and Ferritin

Higher cortisol reactivity was associated with higher IL-6 across pregnancy (β = 0.070, 95% CI [0.007, 0.133], *p* = .029). In trimester-stratified post hoc analyses, this association was significant only in the second trimester (β = 0.118, 95% CI [0.006, 0.230], *p* = .039), and not in the first (β = 0.148, 95% CI [−0.081, 0.378], *p* = .202) or third (β = −0.073, 95% CI [−0.180, 0.034], *p* = .181). CRP was not associated with cortisol reactivity across pregnancy (β = -0.039, 95% CI [−0.123, 0.046], *p* = .365).

Basal cortisol was positively associated with IL-6 centered at 28 weeks (*p* = .039), and a significant basal cortisol × gestational age interaction indicated that higher basal cortisol was associated with a steeper rise in IL-6 across gestation (*p* = .002; for basal cortisol model results, see Supplemental Table 4A). IL-6 was in turn positively associated with ferritin (*p* = .006). After accounting for IL-6, the cortisol × gestational age interaction on ferritin was attenuated and non-significant (*p* = .187), consistent with partial mediation. The indirect effects of basal cortisol on ferritin via IL-6 were estimated at five gestational age windows (12, 20, 28, 34, and 38 weeks; Supplemental Table 4B) and were significant at all windows except 20 weeks (Figure 4). The indirect effect showed a systematic shift across gestation, moving from negative at 12 weeks, through near-zero at 20 weeks, to positive at 28 weeks, 34 weeks, and 38 weeks of gestation.

**Figure 4:**
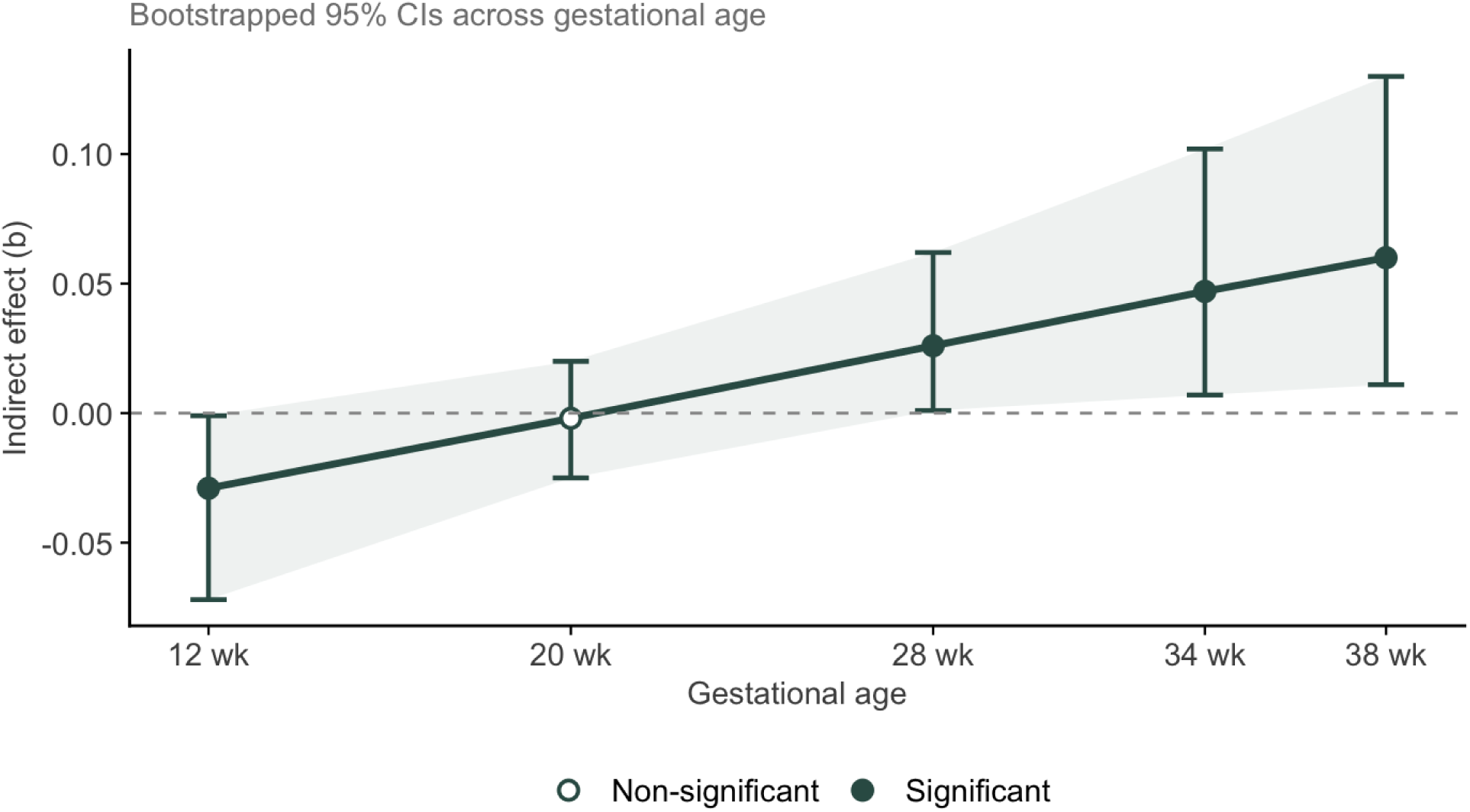
Basal cortisol was associated with ferritin via IL-6 across gestation. The indirect effects of basal cortisol on ferritin via IL-6 were estimated at five gestational age windows (12, 20, 28, 34, and 38 weeks; Supplemental Table 4B) and were significant at all windows except 20 weeks. The indirect effect showed a systematic shift across gestation, moving from negative at 12 weeks (*b* = −0.029, 95% CI [−0.072, -0.001]) through near-zero at 20 weeks (*b* = −0.002, 95% CI [−0.025, 0.02]) to positive at 28 (*b* = 0.026, 95% CI [0.001, 0.062]), 34 (*b* = 0.047, 95% CI [0.007, 0.102]), and 38 weeks (*b* = 0.06, 95% CI [0.011, 0.13]).

CRP was not associated with basal cortisol at 28 weeks (*p* = .639), nor the interaction of basal cortisol with gestational age (*p* = .973; Supplemental Table 4A). CRP was not significantly associated with ferritin, and no significant indirect effects of basal cortisol on ferritin via CRP emerged at any gestational age window. The basal cortisol × gestational age interaction on ferritin remained marginally significant after accounting for CRP (*p* = .087), suggesting CRP does not explain this association. Random effects are summarized in Table 4C.

## DISCUSSION

The present study examined the relationship between cortisol reactivity to a controlled social stressor and iron status across pregnancy in a sample of Black American women living in urban, under resourced environments. Our primary finding was that greater cortisol reactivity to the TSST was associated with lower serum ferritin in the third trimester, an association that persisted after controlling for maternal age, time of day, and concurrent self-reported stress, and was not explained by BMI or RCT group membership in a DHA supplementation trial. This effect was trimester-specific, emerging only in the third trimester, and was not mediated by inflammation. Our secondary finding was that higher basal cortisol was associated with higher IL-6 across gestation, IL-6 was associated with higher ferritin, and IL-6 statistically mediated the association between basal cortisol and ferritin across gestation. Two interpretations warrant consideration, following either a “cumulative depletion hypothesis” or a “HPA dysregulation hypothesis.” Critically, these two interpretations are not mutually exclusive as both may contribute to the trimester-specific pattern observed. To our knowledge, this is the first study to prospectively examine cortisol in relation to iron status across pregnancy, extending prior cross-sectional work linking psychosocial stress to reduced offspring iron status.

The first interpretation is that the trimester-specificity may reflect the cumulative physiological demands of pregnancy. The impact of stress physiology on iron metabolism may only become detectable once maternal iron reserves have been sufficiently depleted through the typical process that accelerates in the third trimester. Decreased ferritin across gestation is thought to reflect the nearly 7-fold increase in iron demand across pregnancy, with a particularly large increase in demand between 24 and 36 weeks of gestation when fetal growth and blood volume expansion is at its greatest (8). In this study, participants’ ferritin levels decreased across pregnancy as expected from prior studies that show both decreasing serum ferritin levels and increased prevalence of iron deficiency across gestation (9,12).

The second interpretation is that cortisol reactivity in the third trimester may carry distinct significance because sustained HPA reactivity late in pregnancy is itself considered atypical. Though basal cortisol increases across gestation, HPA axis reactivity is generally expected to attenuate (30). We found a trend level effect where women with lower basal cortisol showed greater third trimester reactivity across gestation. Women who maintained robust cortisol reactivity despite rising basal levels may represent a subgroup whose HPA axis has not followed the expected pregnancy-related trajectory. Therefore, the overall cortisol pattern inclusive of basal and reactivity measures may be consequential for iron status in late pregnancy, already a state of progressive iron depletion. Speculatively, changes in HPA activation in the third trimester superimposed on this physiological vulnerability may suppress intestinal iron absorption over a sufficient duration to manifest as lower ferritin by the third trimester when cumulative demand is greatest and reserves are most depleted. However, more work is required to disentangle cumulative demand from third trimester reactivity.

### Cortisol and Inflammation

To explore potential mechanisms linking cortisol to iron status, we examined inflammatory markers as possible mediators. Cortisol reactivity was not associated with CRP at any point in pregnancy, and its association with IL-6 was significant only in the second trimester and non-significant (and directionally negative) in the third trimester, when the primary cortisol-ferritin association was observed. These findings are likely related to the chronically elevated inflammatory state observed in this cohort. The mean CRP in the present sample was at or above 9 mg/L across all three trimesters, approaching the threshold of >10 mg/L at which inflammation most meaningfully impacts iron biomarkers in pregnancy (9). In a low-risk, predominantly White, iron-supplemented Irish cohort, McCarthy et al. found that CRP >10 mg/L was associated with significantly higher ferritin and greater iron deficiency compared to women with CRP ≤5 mg/L, consistent with inflammatory iron sequestration. The sustained elevation in our sample across all trimesters suggests a state of chronic inflammation, which may reflect the cumulative physiological burden of chronic psychosocial stress experienced by a marginalized population (24,51). Alternatively, stress-related variation in ferritin due to CRP may have been difficult to detect against an already elevated inflammatory backdrop rather than reflecting a true absence of inflammatory involvement.

The basal cortisol and inflammation findings contrast with the cortisol reactivity and inflammation findings. Although rising basal cortisol, normative in the context of human gestation, was not significantly associated with ferritin across pregnancy, it was associated with a steeper rise in IL-6. IL-6 was in turn positively associated with uncorrected ferritin, consistent with IL-6’s role in upregulating hepcidin which elevates ferritin while reducing circulating iron bioavailability (6,25,27,28). Moreover, we found that the indirect effect of cortisol on ferritin via IL-6 pathways was statistically significant. This pattern raises the possibility that the normative hormonal trajectory of pregnancy might involve inflammatory processes that partially obscure true iron depletion. However, the relatively broad confidence intervals and inconsistency in the direction of effects over the course of pregnancy signals that the findings should be interpreted cautiously, and that additional work is needed to refine iron status assessment in pregnancy.

Hepcidin is hypothesized to be a driver of the link between stress, cortisol, and iron (6). Hepcidin is markedly reduced across the second and third trimesters to facilitate the increased intestinal iron absorption and iron mobilization from stores needed to support maternal red cell mass expansion and fetal iron accrual (8). Chronically elevated hepcidin, driven by sustained low-grade inflammation, would blunt this adaptive suppression to reduce dietary iron absorption over the course of pregnancy and hasten the progressive decline in ferritin that is well underway by the third trimester. In this framework, the cumulative attenuation of iron absorption across a stressed pregnancy may be responsible for lower iron status by the end of gestation. We did not measure hepcidin in this study, so we are unable to directly assess whether cortisol impacted hepcidin in this sample, and the mechanism underlying this association remains to be determined. Pathways beyond IL-6 and CRP warrant consideration in future work, including cortisol-driven effects on intestinal iron absorption and regulatory protein expression as suggested by preclinical work (52,53). For instance, recent research in a mouse model shows that psychological stress increases gut inflammation and that chronically-elevated glucocorticoids promote an inflammatory state, mediated by enteric nervous system glia and neurons (54). Furthermore, there may be hepcidin regulation through pathways beyond IL-6 signaling as demonstrated in murine models (55), and sympathetic nervous system activation as a parallel stress pathway given emerging evidence linking norepinephrine levels to impaired iron transport and status (56).

### Alternative Explanations

An alternative explanation is that low ferritin may itself drive cortisol reactivity. In a randomized trial of pregnant women in rural Nepal, iron and micronutrient supplementation were associated with lower third trimester cortisol and erythropoietin, suggesting that improving iron status may buffer physiological stress responses (57). However, our temporal precedence analyses found that ferritin earlier in the third trimester did not predict cortisol reactivity later within the same individuals, arguing against reverse causation in our sample. This discrepancy may reflect differences in cortisol measurement: prior studies relied on single time-point blood cortisol, which captures a snapshot vulnerable to diurnal variation, physical activity, and transient fluctuation. Our repeated salivary cortisol assessments in response to a controlled laboratory stressor index how the HPA axis functionally responds under stress across gestation, providing a more precise characterization of stress physiology and iron status in pregnancy.

The association between cortisol reactivity and ferritin could reflect stress-related changes in diet or other behaviors that influence iron stores. However, self-reported stress did not predict ferritin in any trimester in our models of cortisol reactivity predicting ferritin. Our findings are consistent with a systematic review that found that cortisol measures more reliably predict adverse birth outcomes than self-reported stress, underscoring the importance of biological indicators in prenatal stress research (58). Moreover, a recent longitudinal study found no evidence that stress and diet quality predict one another across pregnancy (59), making diet an unlikely sole explanation. That said, elevated cortisol responses to acute stressors may influence eating behavior independently of perceived stress, and resource-related stressors not well captured by self-report measures may co-occur with inadequate iron intake. Future work should examine how cortisol, diet, and iron intake co-vary across pregnancy, particularly given that iron deficiency remains prevalent even in low-risk groups with high supplementation rates (9).

### Implications for offspring development

The developing fetus accrues most of its iron in the third trimester to support rapid postnatal growth on an exclusive low-iron diet of human milk before complementary foods are introduced at 6 months (3,8,60–62). Third-trimester maternal iron status is therefore a critical determinant of neonatal iron endowment and infant iron status through at least the first year of life (60,63), making the cortisol–ferritin association identified here a potential upstream risk factor for offspring neurodevelopment (64). The trimester means of maternal ferritin levels observed in this sample, while declining across gestation, were likely not sufficiently low to reduce cord ferritin below the neurologic threshold of <76 μg/L at which neonatal brain iron deficiency and neurodevelopmental compromise are observed (65,66). However, even modestly reduced fetal iron stores at birth accelerate the onset of postnatal iron deficiency, with consequences for brain development (6). Thus, the primary neurodevelopmental risk conferred by stress-related reductions in maternal third-trimester ferritin may operate postnatally (e.g., through earlier exhaustion of neonatal iron stores and a longer duration of deficiency during a period of rapid brain growth) rather than through direct fetal brain iron deprivation *in utero*. This fits preclinical work in non-human primates, which found that maternal stress induced lower iron in offspring postnatally (15). Further, the context of maternal stress may change fetal loading independent of maternal iron status, as recent preclinical work suggests that increased cortisol sampled once during cesarean section and chronic maternal stress in rodents increased placental iron uptake while reducing iron transfer to the fetus (67), consistent with hepcidin-mediated sequestration in placental macrophages under inflammatory conditions. Future work should seek to establish how maternal stress and HPA axis activity directly impacts fetal iron transfer and subsequent neurodevelopment.

**Limitations** Participants showed limited cortisol reactivity to the TSST, with levels highest at arrival and declining across the task. Sufficient time elapsed between visits (mean 5.74–7.50 weeks) to expect renewed cortisol responding to repeated TSST exposure (68,69), suggesting this pattern reflects either chronic stress-related HPA blunting rather than habituation, or reflects an inability of the TSST to trigger an acute cortisol response for the sample as a whole. This is consistent with prior findings in this sample linking discrimination-related stress to reduced cortisol responsiveness (37), and with broader evidence that chronic stress dysregulates HPA functioning in marginalized populations (37,38,41,51,70). As a result, our findings may underestimate the true association between cortisol reactivity and ferritin. We note that maternal stress has been associated with worse neonatal iron stores even at low-to-moderate stress levels, suggesting broader applicability (19).

Hepcidin and soluble transferrin receptor were not measured, precluding direct tests of the cortisol-inflammation-hepcidin pathway. However, ferritin remains a well-validated marker of iron status in pregnancy (9), and their absence does not affect interpretation of the primary findings. Moreover, we were able to adjust and control for inflammation in all ferritin samples, increasing our confidence that the current study reflects an estimate of true iron status. Another limitation is that dietary iron intake and supplement use were also not assessed; however, the socioeconomic homogeneity of the sample suggests participants likely shared similar nutritional risk profiles. Food insecurity, which disproportionately affects low-income populations, is associated with reduced iron intake during pregnancy (71,72).

The sample was homogenous with respect to race, income, and urbanicity, which limits generalizability but also reflects the target population of interest; Black women living in under resourced environments represent a group at elevated risk for both chronic stress and iron deficiency, and understanding stress-iron associations in this population has direct clinical relevance. Black women in the United States face disproportionate rates of gestational iron deficiency and are less likely to be screened or treated for these conditions despite facing more severe clinical consequences (73,74). Understanding how stress physiology contributes to iron status in this population has direct relevance for reducing disparate maternal and infant health outcomes. Although race-specific hemoglobin adjustments have been proposed for Black women, standard WHO pregnancy thresholds were applied given insufficient mechanistic evidence for race-adjusted cutoffs (75). RCT group did not predict cortisol reactivity, ferritin, CRP, or IL-6 at any trimester, indicating that DHA supplementation did not confound primary findings. Taken together, future research should examine cortisol reactivity and iron status across diverse samples with heterogeneous stress exposures, and consider broader measures of stress physiology including hair cortisol, heart rate variability, and total diurnal cortisol output, alongside direct measures of hepcidin, dietary iron, and supplement use.

## CONCLUSION

To our knowledge, this is the first prospective study to examine associations between HPA axis function and iron status across human pregnancy. We demonstrate that greater cortisol reactivity to acute stress predicts lower serum ferritin specifically in the third trimester-- a period of peak fetal iron demand-- and that this association persists independent of self-reported stress and is not explained by the inflammatory markers assessed here. These findings identify maternal stress physiology as a novel upstream contributor to iron deficiency in late pregnancy, with potential consequences for fetal iron accrual and offspring neurodevelopment during a critical window of brain growth. Importantly, the association may be particularly relevant for understanding health disparities in maternal and infant outcomes. Screening for indicators of HPA dysregulation alongside standard iron status assessment may help identify pregnant women at elevated risk for iron deficiency, and stress-reduction interventions during pregnancy warrant investigation as a complementary strategy to supplementation for preventing iron deficiency and its downstream consequences for infant development.

## Data Availability

Data without personal identifiers can be made available upon reasonable request to the authors.

## Acknowledgements

We are grateful to the participants and research assistants from the Nutrition and Pregnancy Study (NAPS) who make this work possible. Thanks are also due to Nandini Kaluvakolanu for running the ferritin assays at the University of Chicago.

## Statement of author’s contributions to the manuscript

KK, KM, and BMR designed research; KK and KM conducted research/curated data; BMR and GFC analyzed data; BMR, GFC, MKG, and KK wrote the paper. BMR had primary responsibility for final content. All authors have read and approved the final manuscript.

## Data sharing plan

Data without personal identifiers can be made available upon reasonable request to the authors.

