## Supplemental Figure 1 for "Cortisol Stress Response is Associated with Iron Status in Pregnancy"

**Supplementary Figure 1.** Participant Flow Diagram.

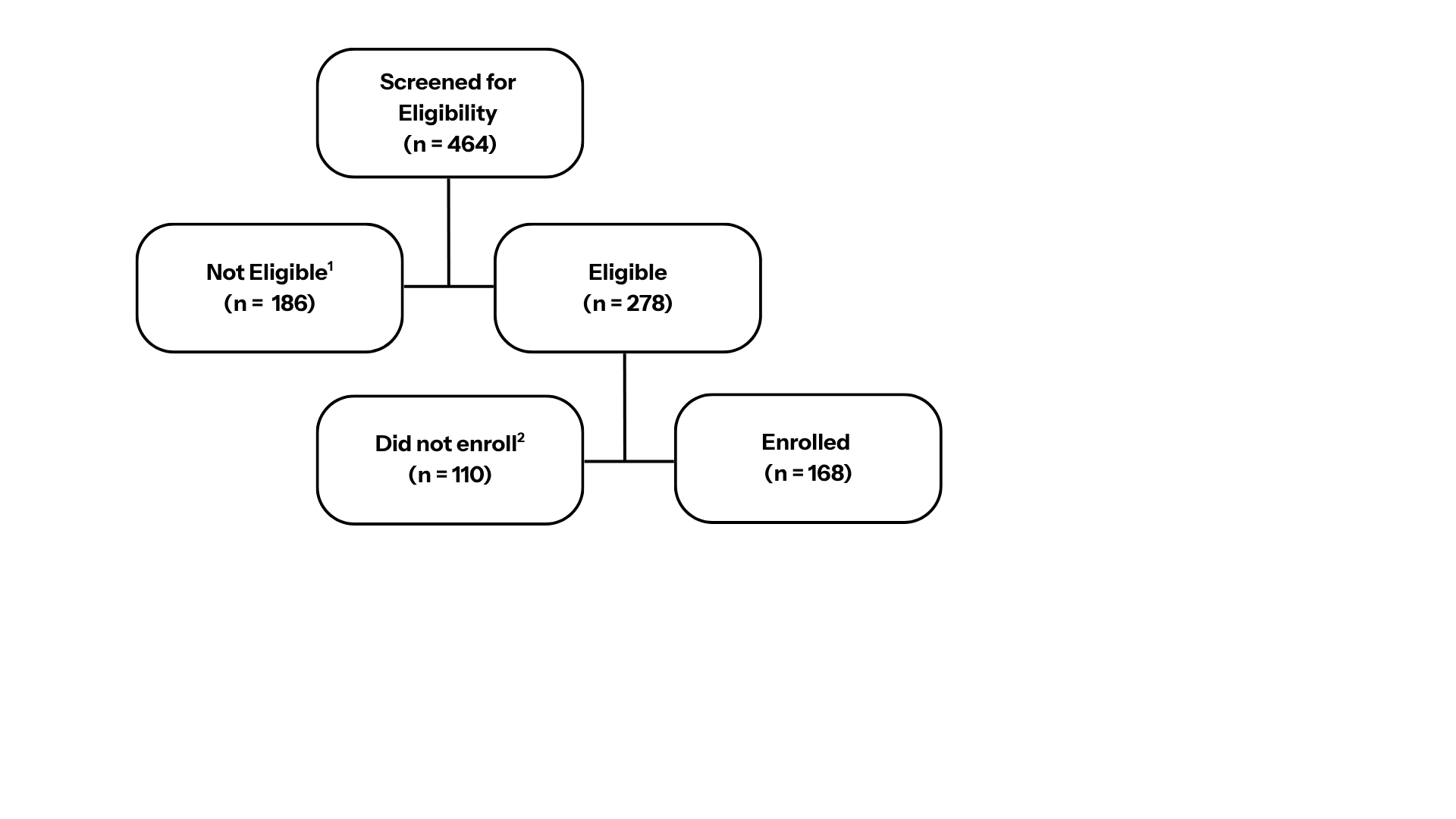

^1^Not eligible after screening (*n* = 186)*:* Gestational Age > 17 weeks (*n* = 40); non-Medicaid insurance (*n* = 25); Exclusionary medical condition (*n* = 22); Substance use (*n* = 19); Adequate fish intake (*n* = 13); non-Black Race (*n* = 13); Maternal age not between 18 and 40 years (*n* = 10); Steroid use (*n* = 7); Allergy to fish, soy, iodine, or strawberry (*n* = 6); Non-partner clinic (*n* = 4); Miscarriage/Termination of pregnancy (*n* = 2); Multiple gestation (*n* = 4); Other Reason (*n* = 22).

^2^Did not enroll after eligibility confirmed (*n* = 110)*:* Gestational age criterion of < 17 weeks exceeded before enrollment (*n* = 84); Declined (*n* = 7); Pregnancy ended (*n* = 5); Other Reason (*n* = 2)
