## Supplemental Text and Tables 1-3 for "Cortisol Stress Response is Associated with Iron Status in Pregnancy"

**Associations between RCT assignment and Trimester-specific hemoglobin levels:** There were no significant differences by RCT group assignment in first trimester hemoglobin levels (*M* = 12.191 vs. 11.675, *t*(24.5) = 1.289, *p*=0.209) or the third trimester hemoglobin levels (*M* = 10.970 vs. 10.722, *t*(52.933) = 0.803, *p*=0.425). However, second trimester hemoglobin levels were significantly higher for those who were randomized to receive DHA supplementation compared to those who did not (*M* = 11.354 vs. 10.758, *t*(34.6)=2.123, *p*=0.041).

**Supplementary Table 1:** *Correlations between variables of interest in the first trimester*

| Variable | *M* | *SD* | 1 | 2 | 3 | 4 | 5 | 6 |
| --- | --- | --- | --- | --- | --- | --- | --- | --- |
| 1. Ferritin (ln) | 4.29 | 0.87 |  |  |  |  |  |  |
| 2. Cortisol Reactivity (ln) | -0.01 | 0.18 | .02 |  |  |  |  |  |
|  |  |  | [-.21, .26] |  |  |  |  |  |
| 3. CRP (ln) | 1.68 | 1.15 | -.11 | -.07 |  |  |  |  |
|  |  |  | [-.34, .13] | [-.29, .16] |  |  |  |  |
| 4. IL-6 (ln) | 7.49 | 0.61 | -.04 | .15 | .62** |  |  |  |
|  |  |  | [-.27, .20] | [-.08, .36] | [.46, .74] |  |  |  |
| 5. Maternal Age (Visit 1) | 25.74 | 4.26 | .15 | -.03 | .05 | .20 |  |  |
|  |  |  | [-.09, .37] | [-.25, .19] | [-.17, .27] | [-.02, .40] |  |  |
| 6. Gestational Age at Visit | 12.00 | 1.24 | -.14 | .08 | .07 | -.00 | -.05 |  |
|  |  |  | [-.37, .09] | [-.15, .29] | [-.16, .29] | [-.23, .22] | [-.26, .17] |  |
| 7. Gestational Age at Birth | 38.53 | 2.30 | -.03 | .07 | -.10 | -.14 | .00 | .08 |
|  |  |  | [-.28, .23] | [-.17, .30] | [-.33, .14] | [-.37, .10] | [-.23, .24] | [-.16, .31] |

*Note.* *M* and *SD* are used to represent mean and standard deviation, respectively. No participants had more than one visit in the first trimester, so all correlations represent one value per participant. Values in square brackets indicate the 95% confidence interval for each correlation. The confidence interval is a plausible range of population correlations that could have caused the sample correlation (Cumming, 2014). * indicates *p* < .05. ** indicates *p* < .01.

**Supplementary Table 2:** *Correlations between variables of interest in the second trimester*

| Variable | *M* | *SD* | 1 | 2 | 3 | 4 | 5 | 6 |
| --- | --- | --- | --- | --- | --- | --- | --- | --- |
| 1. Ferritin (ln) | 3.99 | 0.93 |  |  |  |  |  |  |
| 2. Cortisol Reactivity (ln) | -0.05 | 0.19 | .05 |  |  |  |  |  |
|  |  |  | [-.12, .22] |  |  |  |  |  |
| 3. CRP (ln) | 1.90 | 1.11 | -.03 | .01 |  |  |  |  |
|  |  |  | [-.20, .14] | [-.15, .18] |  |  |  |  |
| 4. IL-6 (ln) | 7.59 | 0.65 | .04 | .09 | .72** |  |  |  |
|  |  |  | [-.13, .21] | [-.08, .25] | [.63, .79] |  |  |  |
| 5. Maternal Age (Visit 1) | 25.79 | 4.21 | -.02 | .00 | .19* | .11 |  |  |
|  |  |  | [-.18, .15] | [-.16, .17] | [.03, .34] | [-.05, .27] |  |  |
| 6. Gestational Age at Visit | 18.97 | 1.92 | -.26** | -.01 | .09 | .05 | .02 |  |
|  |  |  | [-.41, -.10] | [-.17, .16] | [-.07, .25] | [-.12, .21] | [-.14, .18] |  |
| 7. Gestational Age at Birth | 38.61 | 2.55 | .02 | .09 | .03 | .14 | .02 | .02 |
|  |  |  | [-.15, .19] | [-.08, .26] | [-.13, .20] | [-.03, .29] | [-.14, .18] | [-.14, .18] |

*Note.* *M* and *SD* are used to represent mean and standard deviation, respectively. Before computing correlations, mean values of each variable were calculated within-person to account for participants with more than one visit in the second trimester (N = 107). Values in square brackets indicate the 95% confidence interval for each correlation. The confidence interval is a plausible range of population correlations that could have caused the sample correlation (Cumming, 2014). * indicates *p* < .05. ** indicates *p* < .01.

**Supplementary Table 3:** *Correlations between variables of interest in the third trimester*

| Variable | *M* | *SD* | 1 | 2 | 3 | 4 | 5 | 6 |
| --- | --- | --- | --- | --- | --- | --- | --- | --- |
| 1. Ferritin (ln) | 3.44 | 1.09 |  |  |  |  |  |  |
| 2. Cortisol Reactivity (ln) | -0.04 | 0.09 | -.17 |  |  |  |  |  |
|  |  |  | [-.35, .03] |  |  |  |  |  |
| 3. CRP (ln) | 1.73 | 1.15 | .01 | -.15 |  |  |  |  |
|  |  |  | [-.18, .20] | [-.33, .04] |  |  |  |  |
| 4. IL-6 (ln) | 7.70 | 0.58 | .04 | -.15 | .63** |  |  |  |
|  |  |  | [-.15, .22] | [-.32, .04] | [.51, .73] |  |  |  |
| 5. Maternal Age (Visit 1) | 25.89 | 4.22 | -.04 | -.07 | .16 | .14 |  |  |
|  |  |  | [-.22, .15] | [-.25, .12] | [-.02, .33] | [-.03, .31] |  |  |
| 6. Gestational Age at Visit | 31.76 | 1.84 | .07 | -.09 | -.10 | -.11 | .07 |  |
|  |  |  | [-.12, .25] | [-.27, .10] | [-.27, .08] | [-.28, .06] | [-.10, .23] |  |
| 7. Gestational Age at Birth | 38.57 | 2.57 | -.17 | -.09 | .05 | .03 | .02 | .29** |
|  |  |  | [-.35, .02] | [-.27, .10] | [-.13, .22] | [-.14, .21] | [-.14, .19] | [.14, .44] |

*Note.* *M* and *SD* are used to represent mean and standard deviation, respectively. Before computing correlations, mean values of each variable were calculated within-person to account for participants with more than one visit in the third trimester (N = 103). Values in square brackets indicate the 95% confidence interval for each correlation. The confidence interval is a plausible range of population correlations that could have caused the sample correlation (Cumming, 2014). * indicates *p* < .05. ** indicates *p* < .01.
