## Supplemental Table 4 for "Cortisol Stress Response is Associated with Iron Status in Pregnancy"

| **Table 4A.** | | | | | | | | | | |
| --- | --- | --- | --- | --- | --- | --- | --- | --- | --- | --- |
| **Model** | **Parameter** | ***b*** | | ***SE*** | ***t*** | ***df*** | ***p*** | ***B*** | ***95% CI*** | |
| Path C: Cortisol -> Ferritin | Cortisol | 0.041 | | 0.1 | 0.407 | 366.101 | 0.684 | -0.025 | [-0.114, 0.065] | |
| Path C: Cortisol -> Ferritin | Gestational age (centered at 28w) | -0.014 | | 0.015 | -0.91 | 331.265 | 0.363 | -0.293 | [-0.377, 0.210] | |
| Path C: Cortisol -> Ferritin | Group Assignment | -0.263 | | 0.155 | -1.693 | 146.691 | 0.093+ | -0.113 | [-0.245, 0.019] | |
| Path C: Cortisol -> Ferritin | Cortisol x Gestational age | 0.015 | | 0.009 | 1.693 | 328.123 | 0.091+ | 0.066 | [-0.011, 0.142] | |
| ***IL-6 Pathway*** |  |  | |  |  |  |  |  |  | |
| Path A: Cortisol -> IL-6 | Cortisol | 0.109 | | 0.053 | 2.071 | 339.822 | 0.039* | 0.1 | [0.005, 0.196] | |
| Path A: Cortisol -> IL-6 | Gestational age (centered at 28w) | 0.034 | | 0.008 | 4.298 | 318.547 | < .001*** | 0.404 | [0.219, 0.588] | |
| Path A: Cortisol -> IL-6 | Group assignment | 0.042 | | 0.101 | 0.418 | 156.705 | 0.676 | 0.03 | [-0.110, 0.170] | |
| Path A: Cortisol -> IL-6 | Cortisol x Gestational age | 0.015 | | 0.005 | 3.196 | 315.038 | 0.002** | 0.173 | [0.066, 0.279] | |
| Path B: IL-6 -> Ferritin | IL-6 | 0.236 | | 0.085 | 2.779 | 408.376 | 0.006** | 0.143 | [0.042, 0.245] | |
| Path B: IL-6 -> Ferritin | Cortisol | 0.017 | | 0.099 | 0.175 | 365.841 | 0.861 | 0.01 | [-0.100, 0.119] | |
| Path B: IL-6 -> Ferritin | Gestational age (centered at 28w) | -0.022 | | 0.015 | -1.415 | 333.318 | 0.158 | -0.157 | [-0.374, 0.061] | |
| Path B: IL-6 -> Ferritin | Group assignment | -0.278 | | 0.156 | -1.778 | 144.316 | 0.078+ | -0.119 | [-0.251, 0.013] | |
| Path B: IL-6 -> Ferritin | Cortisol x Gestational age | 0.012 | | 0.009 | 1.323 | 328.347 | 0.187 | 0.084 | [-0.041, 0.208] | |
| ***CRP Pathway*** |  |  | |  |  |  |  |  |  | |
| Path A: Cortisol -> CRP | Cortisol | 0.032 | | 0.069 | 0.47 | 302.993 | 0.639 | 0.017 | [-0.054, 0.088] | |
| Path A: Cortisol -> CRP | Gestational age (centered at 28w) | -0.01 | | 0.01 | -0.973 | 294.553 | 0.331 | -0.067 | [-0.204, 0.069] | |
| Path A: Cortisol -> CRP | Group assignment | 0.076 | | 0.193 | 0.395 | 154.865 | 0.693 | 0.031 | [-0.122, 0.184] | |
| Path A: Cortisol -> CRP | Cortisol x Gestational age | 0 | | 0.006 | -0.033 | 291.979 | 0.973 | -0.001 | [-0.079, 0.077] | |
| Path B: CRP -> Ferritin | CRP | 0.069 | | 0.053 | 1.284 | 300.617 | 0.2 | 0.073 | [-0.039, 0.184] | |
| Path B: CRP -> Ferritin | Cortisol | 0.043 | | 0.1 | 0.429 | 362.08 | 0.668 | 0.024 | [-0.086, 0.134] | |
| Path B: CRP -> Ferritin | Gestational age (centered at 28w) | -0.013 | | 0.015 | -0.867 | 328.754 | 0.387 | -0.095 | [-0.310, 0.120] | |
| Path B: CRP -> Ferritin | Group assignment | -0.269 | | 0.156 | -1.724 | 144.608 | 0.087+ | -0.116 | [-0.248, 0.016] | |
| Path B: CRP -> Ferritin | Cortisol x Gestational age | 0.015 | | 0.009 | 1.718 | 324.923 | 0.087+ | 0.108 | [-0.016, 0.232] | |
| **Table 4B.** | | | | | |  |  |  |  | |
| **Pathway** | **Gestational Age Group** | ***b*** | ***95% CI*** | | | | | | | **Significance** |
| Via IL-6 | 12 weeks | -0.029 | [-0.072, -0.001] | | | | | | | Significant |
| Via IL-6 | 20 weeks | -0.002 | [-0.025, 0.020] | | | | | | | N.S. |
| Via IL-6 | 28 weeks | 0.026 | [0.001, 0.062] | | | | | | | Significant |
| Via IL-6 | 34 weeks | 0.047 | [0.007, 0.102] | | | | | | | Significant |
| Via IL-6 | 38 weeks | 0.06 | [0.011, 0.130] | | | | | | | Significant |
| Via CRP | 12 weeks | 0.002 | [-0.011, 0.020] | | | | | | | N.S. |
| Via CRP | 20 weeks | 0.002 | [-0.007, 0.015] | | | | | | | N.S. |
| Via CRP | 28 weeks | 0.002 | [-0.009, 0.017] | | | | | | | N.S. |
| Via CRP | 34 weeks | 0.002 | [-0.014, 0.021] | | | | | | | N.S. |
| Via CRP | 38 weeks | 0.002 | [-0.018, 0.025] | | | | | | | N.S. |
| **Table 4C.** | | | | |  |  |  |  |  | |
| **Model** | **Between Person Variance** | **Residual Variance** | **ICC** | | | | | | |  |
| Path A: Cortisol -> IL-6 | 0.287 | 0.144 | 0.666 | | | | | | |  |
| Path A: Cortisol -> CRP | 1.175 | 0.229 | 0.837 | | | | | | |  |
| Path B: IL-6 -> Ferritin | 0.608 | 0.537 | 0.531 | | | | | | |  |
| Path B: CRP -> Ferritin | 0.605 | 0.549 | 0.524 | | | | | | |  |
